# Stochastic Modeling of Intra- and Inter-Hospital Transmission in Middle East Respiratory Syndrome Outbreak

**DOI:** 10.1101/2024.07.17.24310554

**Authors:** Youngsuk Ko, Jacob Lee, Eunok Jung

## Abstract

Middle East Respiratory Syndrome (MERS) is an endemic disease that presents a significant global health challenge characterized by a high risk of transmission within healthcare settings. Understanding both intra- and inter-hospital spread of MERS is crucial for effective disease control and prevention. This study utilized stochastic modeling simulations to capture inherent randomness and unpredictability in disease transmission. This approach provides a comprehensive understanding of potential future MERS outbreaks under various scenarios in Korea.

Our simulation results revealed a broad distribution of case number, with a mean of 70 and a credible interval of [0, 315]. Additionally, we assessed the risks associated with delayed outbreak detection and investigated the preventive impact of mask mandates within hospitals. Our findings emphasized the critical role of early detection and the implementation of preventive measures in curbing the spread of infectious diseases. Specifically, even under the worst-case scenario of late detection, if mask mandates achieve a reduction effect exceeding 55%, the peak number of isolated cases would remain below 50.

The findings derived from this study offer valuable guidance for policy decisions and healthcare practices, ultimately contributing to the mitigation of future outbreaks. Our research underscores the critical role of mathematical modeling in comprehending and predicting disease dynamics, thereby enhancing ongoing efforts to prepare for and respond to MERS or other comparable infectious diseases.

**Author summary:** In our study, we used stochastic modeling simulations to understand the potential future outbreaks of Middle East Respiratory Syndrome (MERS) in Korea. Our simulations revealed a wide range of possible case numbers, emphasizing the unpredictable nature of disease transmission. We found that early detection of an outbreak and the implementation of preventive measures, such as mask mandates in hospitals, play a critical role in controlling the spread of infectious diseases. Even in the worst-case scenario of late detection, if masks are mandated and achieve a reduction effect exceeding 55%, the peak number of isolated cases would remain below 50. Our research highlights the importance of mathematical modeling in understanding and predicting disease dynamics, providing valuable insights for policy decisions and healthcare practices. This work contributes to the ongoing efforts to prepare for and respond to MERS or other similar infectious diseases.

## Introduction

Middle East respiratory syndrome (MERS) results from infection with the Middle East respiratory syndrome coronavirus (MERS-CoV) [1], a zoonotic pathogen capable of transmission between animals and humans. Clinically, MERS patients exhibit varying degrees of symptoms, including fever, cough, and shortness of breath [2]. Disease progression leads to complications such as pneumonia, acute respiratory distress syndrome, kidney failure, and multiorgan dysfunction. The average fatality rate stands at approximately 36%, with regional variations ranging from 14% to 44% [3–5] based on reported cases. MERS was initially identified in Saudi Arabia in 2012 [2] and continues to sporadically emerge in endemic regions, notably Saudi Arabia, which reports the highest case count [5]. As of 2024, Saudi Arabia has documented 2,204 cases, resulting in 862 deaths [5].

Subsequent to Saudi Arabia, South Korea reported the second highest incidence of MERS infections, predominantly linked to a singular outbreak. In 2015, Korea faced a MERS outbreak originating from an individual who had recently visited the Middle East [6, 7]. The disease primarily disseminated within hospital settings, resulting in 186 confirmed cases and 36 fatalities—a significant public health challenge for the country [8]. Notably, this outbreak exhibited a pronounced risk of transmission within hospitals, while general community spread remained limited [9, 10]. These dynamics emphasize the critical need to comprehend both intra- and inter-hospital MERS transmission pathways for effective disease management and prevention.

Mathematical modeling plays a crucial role in investigating infectious diseases, offering a structure approach for deciphering and forecasting transmission dynamics [11]. Key advantages of such models lie in their ability to yield quantitative insights. By capturing intricate interactions among hosts, pathogens, and the environment, these models facilitate outbreak prediction, intervention assessment, and informed public health policy. Unlike decisions based on general observations, these models rely on precise numerical data. Policymakers benefit from these accurate data when evaluating potential interventions, such as vaccination campaigns, travel restrictions, and enforcing social distancing measures [12–14]. Specifically for MERS, mathematical models have been pivotal in uncovering transmission patterns, predicting outbreaks, and identifying effective public health measures [15, 16]. Assessments have highlighted the potential for sustained MERS transmission, emphasizing hospital-based spread [17, 18]. Investigating superspreading events relative to the basic reproductive number has been pivotal [19, 20]. Agent-based modeling techniques illuminate the impact of superspreading events on the MERS transmission dynamics of MERS, particularly during the Korean outbreak [21].

In this study, we utilize stochastic modeling simulations to capture the inherent randomness and unpredictability of disease spread, encompassing both intra- and inter-hospital transmission dynamics. Our model yields a comprehensive understanding of future MERS outbreaks across diverse scenarios, informing effective control strategies in South Korea. Additionally, we assess the risk associated with delayed outbreak detection and investigate the preventive impact of mask mandates within healthcare facilities. These considerations are important given the current global health landscape, where timely detection and preventive measures are essential in managing infectious disease spread. Our research aims to contribute to preparedness and response efforts for MERS and related infectious diseases, offering valuable insights to guide policy decisions and healthcare practices, ultimately mitigating of future outbreaks.

## Materials and methods

### Stochastic modeling of MERS outbreak

Drawing upon the Susceptible-Exposed-Infectious-Recovered (SEIR) mathematical model, we constructed a framework that accounted for both intra- and inter-hospital transmission of MERS. Our investigation focused on Gangnam District in Seoul, Korea, which comprises two tertiary referral hospitals (average bed count: 1,533), two general hospitals (average bed count: 225), and 32 smaller hospitals (average bed count: 73) [22]. The local community population stands at 650,000 [23]. Notably, our analysis excluded smaller clinics. Within our model, hosts traverse six epidemiological stages: susceptible (*S*), exposed (*E*), infectious (*I*), hospitalized (*H*), isolated (*Q*), and recovered (*R*). In the hospital context, hosts are further categorized into medical staff (subscript *M*), inpatients (*P*), and visitors (*V*). Importantly, we did not differentiate superspreaders in this study. Figure 1 depicts the flowchart of our epidemiological model, illustrating disease progression stages. Solid arrows denote disease transmission pathways. The terms 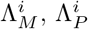, and 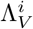 represent the force of infection for nosocomial infections, influenced by 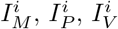, and *H*^*i*^. Here, the superscript *i* denotes hospital identifiers. Additionally, Λ represents the force of infection for local spread, with contributions from *I*, 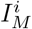, and 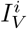. We assume a Markovian process for disease transmission, where future states depend solely on the current state, independent of preceding events. Dashed arrows with numerical labels indicate delayed reactions (non-Markovian process) in disease progression. To simulate our model, we aggregate recorded time delays and incorporated them into the simulation [24]. Transmission rates and reductions in intra-hospital transmission due to non-pharmaceutical interventions (NPIs) are estimated based on the Pyeongtaek St. Mary’s Hospital outbreak, the site of the initial hospital cluster infection [25]. Detailed description of model formulation, fitting distributions of time delays, and parameter estimation is included in S1 Text.

**Fig 1.**
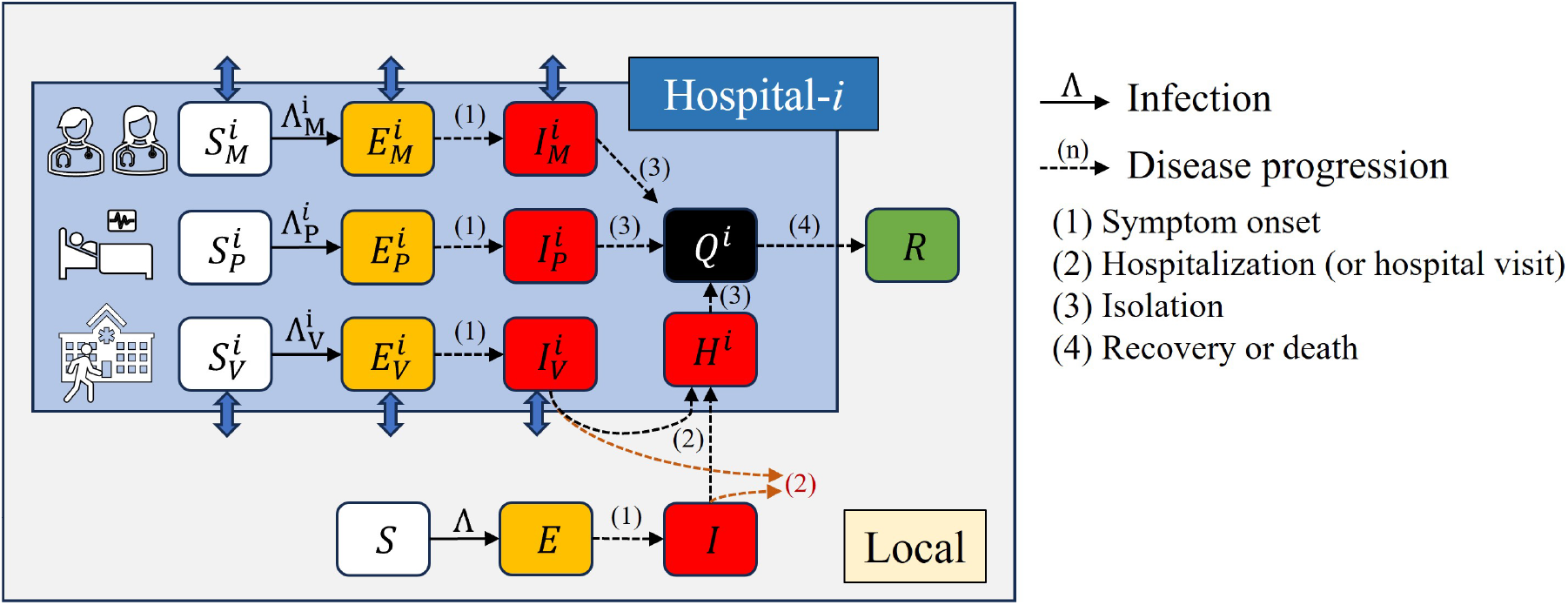
Flow diagram of Middle East Respiratory Syndrome transmission model considering intra- and inter-hospital transmission.

### Model simulation scenarios

For the baseline scenario, our model simulation commences with the introduction of a single primary case into the local community. To align with the 2015 situation in Korea, outbreak detection and reactive NPIs are initiated five days after the index case admission [8]. Upon symptom onset, a host is randomly admitted to a hospital. Figure 2 shows the simulation flow, encompassing outbreak recognition and intervention application. Once the outbreak is identified, we implement NPIs: hospital visits are prohibited (i.e., *V*_*i*_ transitions to susceptible (*S*)), and the intra-hospital transmission rates decrease by 59%. The estimation of this reduction is detailed in S1 Text. Additionally, beyond the baseline scenario, we explore additional scenarios considering the following factors:

**Fig 2.**
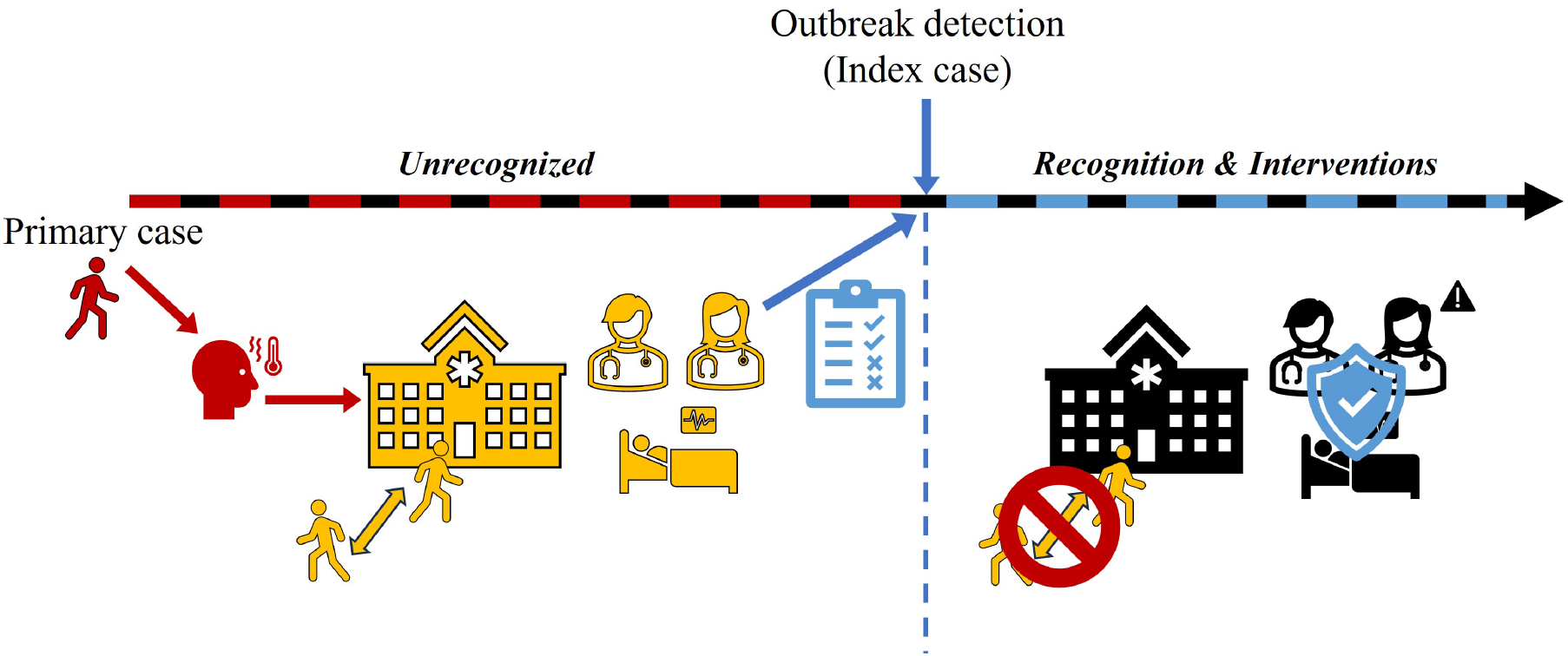
Flow of the simulation from the import of the primary case to the outbreak recognition and reactive interventions.

- Delay in outbreak detection: The risk associated with outbreak recognition delay is investigated, considering a range from 1 to 28 days. Notably, for the baseline scenario, this delay is specifically quantified as five days.
- Mask mandates in hospital: In hospital settings, the preventive impact of mask-wearing intervention is assessed accounting for mask type and enforcement. The resulting transmission reduction effect ranges from 0% (baseline scenario) to a maximum of 80%.

## Results

### Baseline scenario simulation and sensitivity analysis

In Figure 3, panels A, B, and C depict the distributions of confirmed cases, exposed hospitals, and outbreak duration, respectively. We define an exposed hospital that has at least one infected host (*E, I, H*), and outbreak duration is the time from onset to the isolation of the last infected host. Among the simulation runs, approximately 8% conclude without secondary infections. The mean number (95% credible interval; CrI) for confirmed cases, exposed hospitals, and outbreak duration are 70 ([0,315]), 3 ([1,12]), and 119 ([33,253]), respectively. Panel D presents a box-chart graph combining panels A and B, with a red asterisk marking the MERS outbreak in Korea in 2015 (case number: 186, exposed hospitals: 8). Notably, a strong correlation exists between confirmed cases and exposed hospitals (coefficient: 0.70, p-value approximately zero). Similarly, panel E presents a scatter plot depicting the relationship between the number of confirmed cases and outbreak duration. The color scale indicates clustering, with samples closer to yellow representing stronger clustering. A correlation analysis yields a coefficient of 0.77, smaller than the previously estimated coefficient. Additionally, the associated p-value is close to 0.

**Fig 3.**
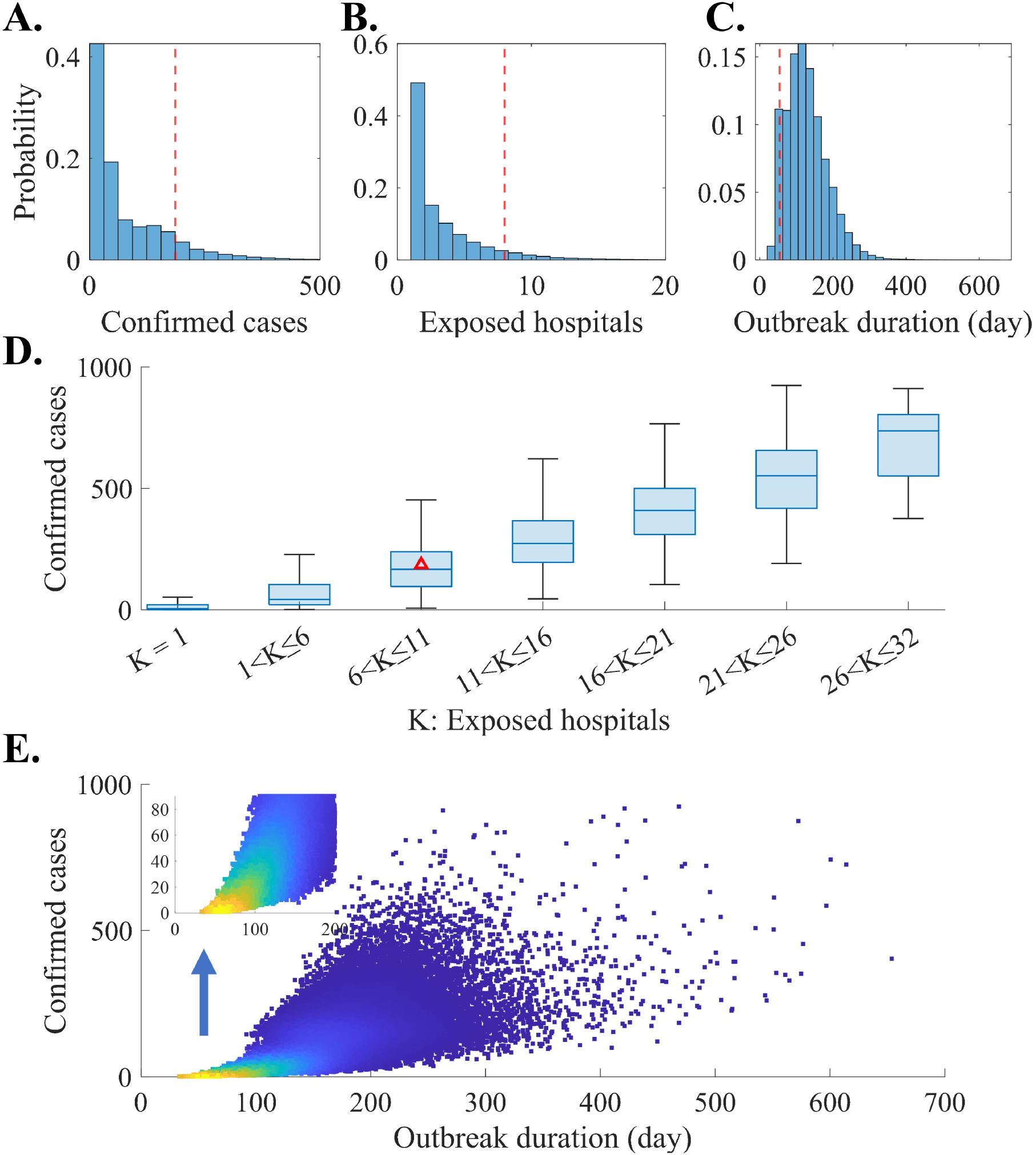
Baseline simulation results of outbreak outcomes: Distribution of the number of confirmed cases, exposed hospitals, and outbreak duration (A, B, and C), Distribution of confirmed cases using a box-chart graph according to the range of exposed hospitals (D), Weighted scatter plot of the number of confirmed cases according to the outbreak duration (E). From panels A to C, red dashed vertical lines indicate the actual outbreak outcome during the 2015 MERS outbreak in Korea, which is also marked using red triangle in a panel D.

Figure 4 shows the time series graphs of infected hosts. Panels A, B, and C correspond to the number of hosts in the exposed, infectious, and isolated states, respectively. The black curve represents the mean value, whereas the red area indicates the 95% CrI. Approximately 60 days after the initial onset, the exposed and infectious stages peak mean values of 6 and 4, with 95% CrI of [0,29] and [0,22], respectively. Notably, the number of isolated patients could exceed 100 considering the 95% CrI.

**Fig 4.**
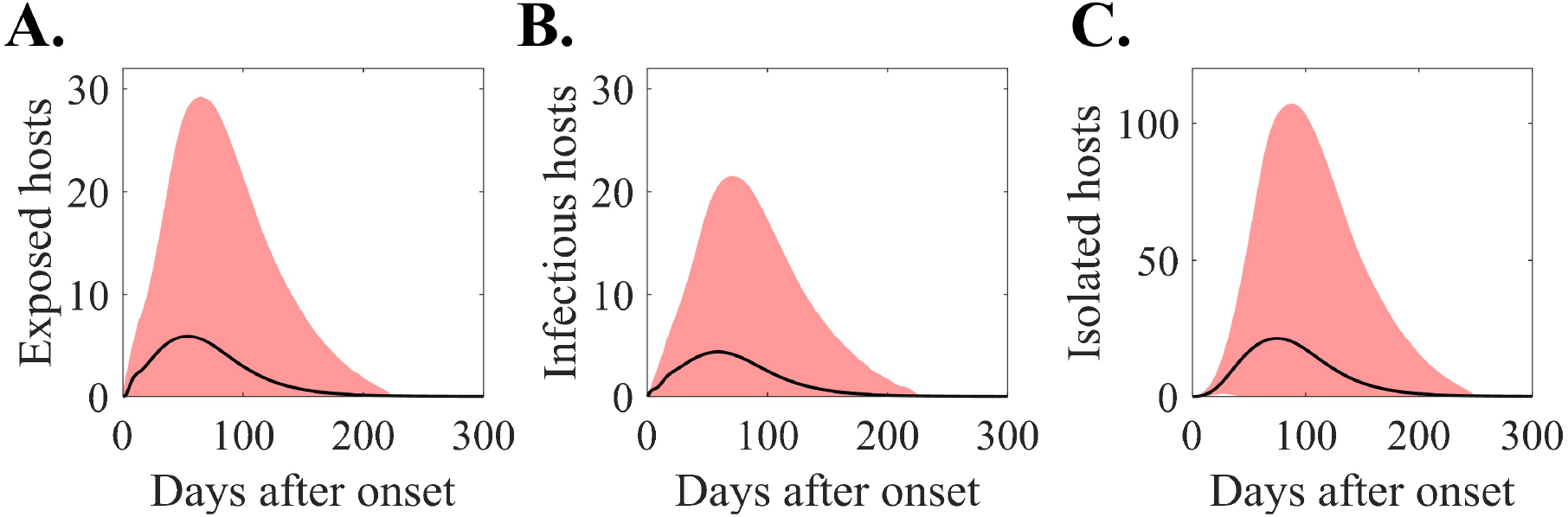
Number of infected hosts in different stages: Disease-exposed (A), Infectious (B), Isolated (C).

Furthermore, we performed a sensitivity analysis to identify the key factors influencing the outbreak. Using Latin Hypercube Sampling, we quantified the Partial Rank Correlation Coefficient (PRCC) values [26]. Our model incorporated several input parameters: outbreak detection timing (*τ*_*rec*_), intra-hospital transmission rates (*β*_*H*_), intra-hospital infectious period for the primary case 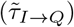, intra-hospital infectious period excluding the primary case (*τ*_*I*→*Q*_), local transmission rate (*β*_*L*_), local infectious period for the primary case 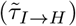, local infectious period excluding the primary case (*τ*_*I*→*H*_), and incubation period (*τ*_*E*→*I*_). We performed 200,000 simulation runs, introducing a uniform variation of *±*30% based on default baseline values. Notably, *β*_*H*_ encompassed all transmission rates within the hospital. Additionally, we accounted for delay-inputs by adjusting the variation ratio using values generated during the model simulation. The output of the model represented the cumulative number of infections, capturing the transition from susceptible to exposed stage.

Figure 5 illustrates the time-dependent PRCC values for each input. *β*_*H*_ consistently exhibits the highest PRCC, ranging from 0.16 to 0.31 throughout the observation period. Following closely are 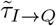 (PRCC: [0.07, 0.12]) and *τ*_*I*→*Q*_ (PRCC: [0.01, 0.21]). During the initial 15 days after onset, 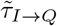 exhibits higher PRCC; however, *τ*_*I*→*Q*_ surpasses it subsequently. The parameter *β*_*L*_ has the second-highest PRCC values within the range [0.04, 0.08]. Notably, 87 days after onset, the PRCC of *β*_*L*_ exceeds that of 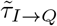. Interestingly, *τ*_*E*→*I*_ is the sole input among all factors where the sign of PRCC changes from positive to negative. Initially positive (maximum 0.03), PRCC transitions to negative (minimum -0.02) 13 days after the outbreak began.

**Fig 5.**
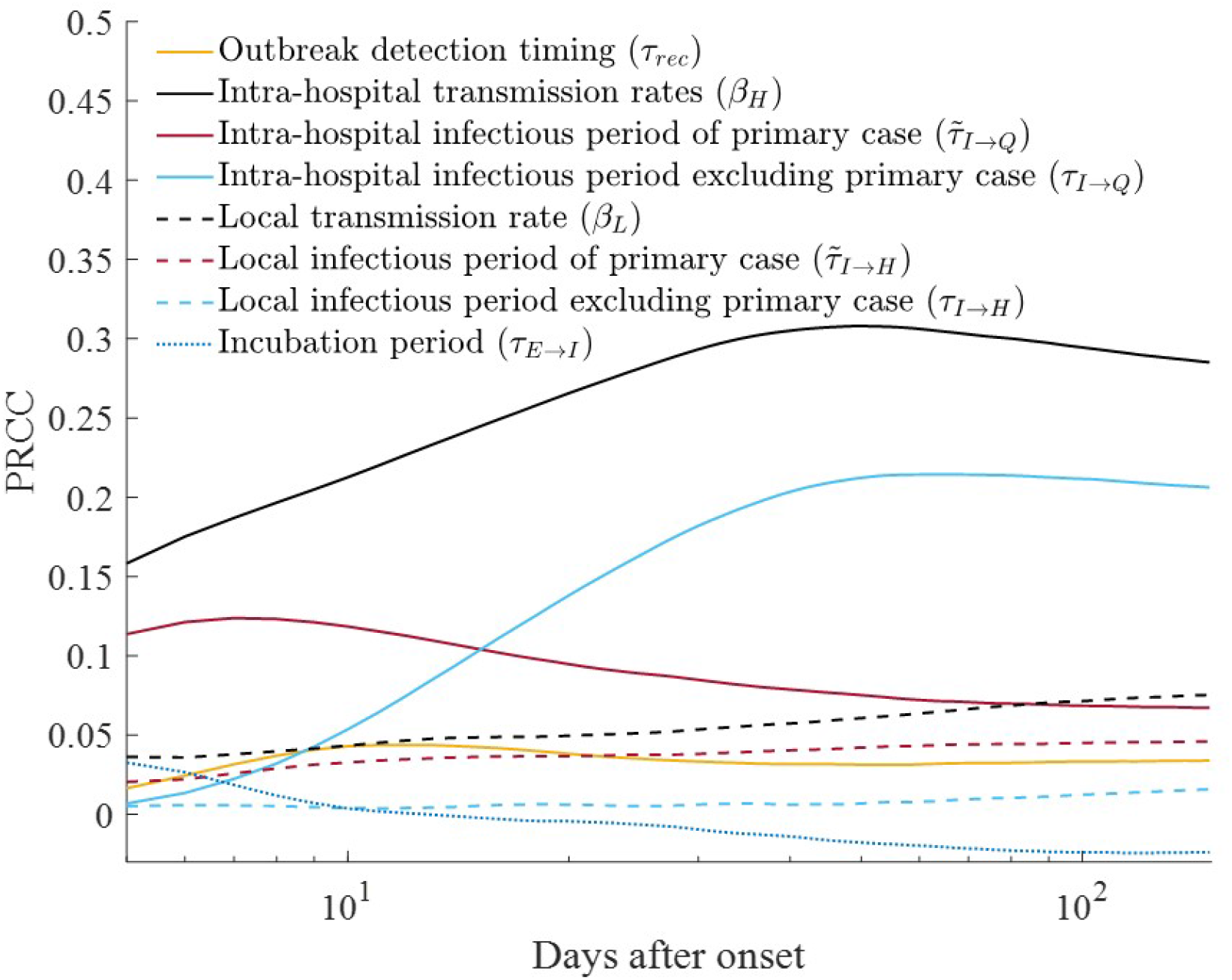
Results from the parameter sensitivity analysis, quantified using the Partial Rank Correlation Coefficient (PRCC). Notably, the X-axis (time, days after onset) is log-scaled.

### Risk of late detection and preventive impact of mask-wearing in hospital

We investigated the impact of delayed outbreak detection and preventive effects of mask mandates within hospitals during an outbreak. Specifically, we analyzed the peak number of isolated patients —a critical risk indicator—by simultaneously adjusting two factors. Figure 6 illustrates the results using filled contour plots, depicting the mean value (panel A) and the maximum value within the 95% CrI (panel B). When mask mandates have no effect (represented by a value of 0 on the X-axis), the peak number of isolated patients can exceed 350 within the 95% CrI if outbreak detection is delayed by up to 28 days. However, with the fastest detection (one day), the peak is expected to average approximately 25 and reach up to 125 within the 95% CrI. When mask mandates have an effect exceeding 40%, the peak number of isolated patients—considering outbreak detection at its latest (28 days)—is smaller than the peak without mask mandates when comparing maximum values within the 95% CrI. Furthermore, if the effect of mask mandates exceeds 70%, the maximum number of peak isolated patients within the 95% CrI under late outbreak detection, is smaller than the mean when detection is fastest without mask mandates. The contour line representing the mean peak number of isolated patients at 50 is of particular significance. Without the effect of mask mandates, this threshold is achieved with a 12-day detection delay. When extrapolated to the far right, the contour line corresponds to a scenario involving a 28-day detection delay and a 30% impact from mask mandates.

**Fig 6.**
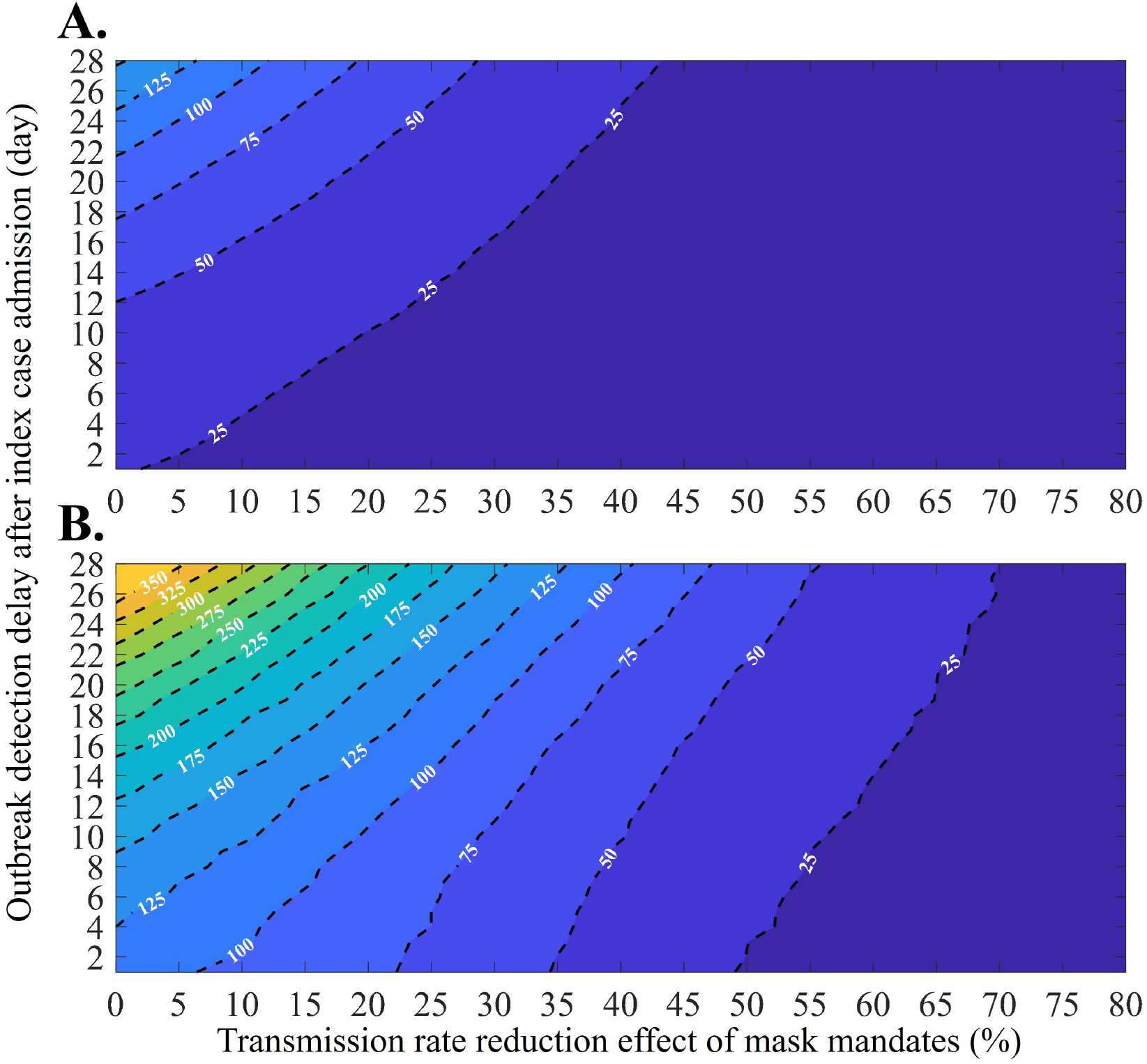
Filled contour plots depict the peak number of isolated patients as a function of changes in the transmission reduction effect due to mask mandates and outbreak detection delay: Mean value (A), and Maximum value within the 95% CrI (B).

## Discussion

Contrary to observed clusters in Korea and other locations, MERS did not significantly spread within general local communities [6]. However, transmission risk was notably high in specific settings, such as hospitals [7, 9, 10]. To address this phenomenon, we conducted a scenario-based study using stochastic modeling to assess the uncertainty and potential spread risk associated with the importation of MERS or similar diseases.

The baseline scenario simulation results revealed a broad distribution of case numbers, ranging from 0 to 315 within the 95% CrI. Additionally, the simulation indicated a high probability of small-scale outbreaks (Figure 3A). This phenomenon arises because an outbreak initiated by a single case is significantly influenced by the individual events, potentially leading to early containment or limited transmission to the second or third generation of infection. Notably, the number of confirmed cases observed during the 2015 Korean fell slightly below the upper bound of the CrI for the baseline scenario (upper 10%). Furthermore, when we further analyzed the simulation results by considering the number of hospitals exposed to the disease, the distribution of confirmed cases could vary (Figure 3D). In the 2015 simulation results, which align with the observed number of exposed hospitals (nine hospitals), the case represents the upper 44% of the distribution. Therefore, we can conclude that the 2015 scenario is not an unrealistic situation. Additionally, Figure 4 illustrates the estimated number of infected hosts at a specific time. This information serve as a reference for establishing tracking targets during contact tracing, particularly for epidemiological investigators, at the outset of an outbreak. For instance, 20 days after the primary case onset, the mean number of exposed and infectious hosts is 3 and 2 (with 95% CrI [0,10] and [0,7]), respectively.

The sensitivity analysis conducted on the baseline scenario yielded quantitative insights (Figure 5). Notably, the infectious period of the initial primary case was found to be more significant than that of the remaining infected individuals during the early phase of the outbreak, as revealed by the time-varying PRCC. In our investigation, we noted that the local community’s transmission rate significantly influenced the outbreak scale. This observation raises the question of whether NPIs are also warranted at the local community level. However, considering the socio-economic burden associated with implementing NPIs locally —a different scale from the hospital-level control measures—the results of our sensitivity analysis paradoxically underscores the importance of early detection of the index case (primary case) rather than immediate NPI application within the local community. Notably, during the COVID-19 pandemic, several countries, including Korea, faced a substantial increase in NPI burden when the outbreak reached a nationwide scale [27, 28].

Our simulation results reveal that rapid detection within hospitals can significantly mitigate outbreak scale, while delayed detection exacerbates the situation (Figure 6B). The peak size of isolated patients emerges as a critical factor affecting medical resources; exceeding local community capacity can lead to rapid deterioration [29]. In Gangnam district (2024), where 77 isolation beds are available [22], a detection delay of more than 18 days —without preventive mask mandates —could result in medical collapse. Our findings underscore the need for a constant level of isolation ward capacity and provide a realistic risk assessment (.g., a four-day detection delay after index admission corresponds to an upper-bound estimate of approximately 125 cases within the 95% CrI).

In scenarios where a primary case is introduced into a local community, rapid outbreak detection poses challenges. Such cases can be misdiagnosed as other diseases not requiring isolation, and the testing process itself consumes time. Consequently, declaring an outbreak immediately upon case admission is realistically difficult [30, 31]. However, our study reveals that maintaining mask mandates can have a substantial initial outbreak-blocking effect, even when these measures are implemented within hospitals rather than being mandatory at the community level. While a single day of outbreak detection delay poses a risk, mask mandates exhibit an effect similar to advancing outbreak detection by several days. For instance, if hospital mask mandates achieves a 50% preventive effect (Figure 6), the peak size of isolated patients remains below 75, even within the maximum range of the 95% CrI. Conversely, without mask mandates, an outbreak recognized a day after index case admission results in more than 100 peak isolated patients within the 95% CrI. In summary, our findings suggest that implementing masks within hospitals can alleviate the burden of maintaining community isolation beds.

Despite the impact of the COVID-19 pandemic, mandatory mask-wearing in Korean hospitals ceased in 2024 [32]. This change was motivated by concerns related to discomfort and health issues faced by workers and patients. However, considering the potential benefits during outbreaks of new infectious disease, policy decisions based on indirect early detection methods (such as sewage-based data from neighboring countries or those with frequent exchanges) may be necessary for implementing mask mandates.

The methodology of this study accommodates straightforward parameter adjustments, enabling analysis across diverse diseases context, scenarios, and varying national and healthcare system environments. Specifically, parameters —such as community transmission rates, incubation periods, and infectious periods —vary based disease characteristics and medical environment. Consequently, the framework of this study is poised to serve as a valuable tool for anticipating future emerging infectious diseases.

The limitations of this study are as follows: (1) The transmission rate within the hospital can be heterogeneous depending on the hospital environment; however, in this study, the parameters estimated from the Pyeongtaek St. Mary’s Hospital case were applied uniformly across all hospitals. (2) Unreported cases or superspreaders could not be distinguished (3) The phenomenon of doctor shopping, where the same patient visits multiple hospitals, was not considered [33]. (4) Patients transfer between hospitals and maximum isolation spaces capacity were also omitted. Future studies should address these limitations.

## Data Availability

All data produced in the present study are available upon reasonable request to the authors

## Supporting information

### S1 Text. Supplementary appendix

This contains detailed explanation of methods for the research.

## Acknowledgments

This research was supported by the Government-wide R&D Fund Project for Infectious Disease Research (GFID), Republic of Korea (grant No. HG23C1629). This paper is supported by the Korea National Research Foundation (NRF) grant funded by the Korean government (MEST) (NRF-2021R1A2C100448711). The funders had no role in study design, data collection and analysis, decision to publish, or preparation of the manuscript.

