## Supplementary material for "Stochastic Modeling of Intra- and Inter-Hospital Transmission in Middle East Respiratory Syndrome Outbreak": S1 text

#### 1 Model formulation

Drawing upon an SEIR -type mathematical model, we formulated a comprehensive framework considering intra- and inter-hospital disease transmission. Within this model, we delineated six distinct epidemiological stages relevant to disease propagation: susceptible ($S$), exposed ($E$), infectious ($I$), hospitalized ($H$), isolated ($Q$), and recovered ($R$). Specifically, within the hospital context, hosts were further categorized into sub-groups: medical staff (denoted by subscript $M$), inpatients ($P$), and visitors ($V$). Notably, we did not differentiate superspreaders in this study. The flowchart depicted in Fig 1 visually represents the epidemiological process captured by our model. The superscript $i$ denotes the hospital identifier. Solid lines represent infection transmission events following a Markovian process that disregards past events. The remaining dotted lines correspond to non-Markovian processes, considering past events, such as the incubation period after infection exposure (1, $\tau_{E\to I}$), the infectious period of visitors and local community hosts (2, $\tau_{I\to H}$), intra-hospital transmission period (3, $\tau_{I\to Q}$ and $\tau_{H\to Q}$), and isolation treatment duration (4, $\tau_{Q\to R}$). In this study, we estimated these delays using gamma distributions based on data from confirmed patients and the MATLAB built-in function fitdist. These estimates were incorporated into the model simulation [1]. Fig 2 illustrates the fitted time delay distribution. We assumed a fixed isolation period of 28 days, while other time delays were generated from the following fitted distributions:

$$\tau_{E\to I} \sim \Gamma(4.4493, 1.5702),$$
$$\tau_{I\to H} = \tau_{H\to Q} \sim \Gamma(2.8118, 1.7996).$$

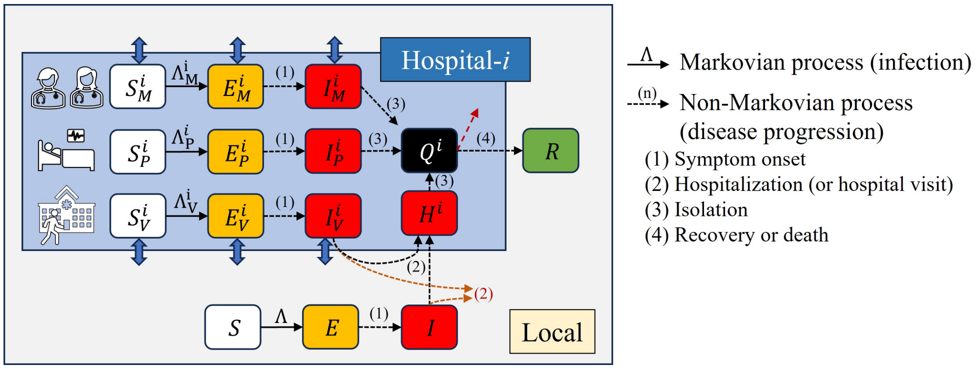


**Fig 1.** Flow diagram of Middle East Respiratory Syndrome outbreak model considering intra- and inter-hospital transmission.

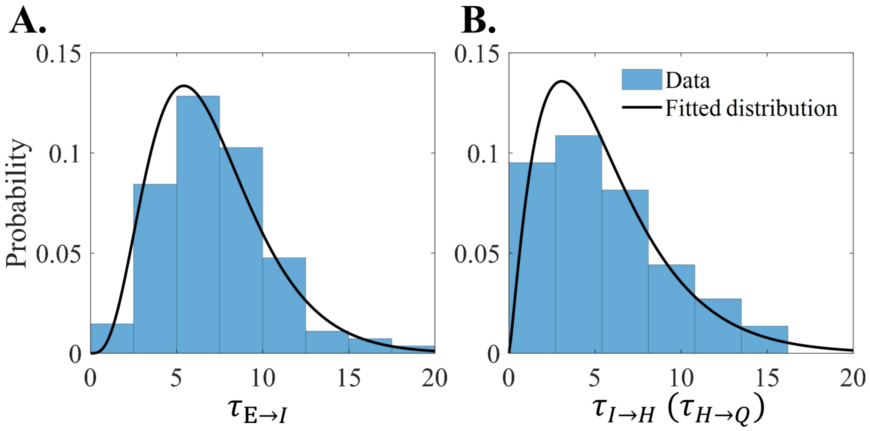


**Fig 2.** Distribution of time delay and fitted curve. Incubation period(A), Infectious period (B).

Both visitors and medical staff can transmit infections beyond the hospital premises. If visitors become infected, they are randomly assigned to one of the hospitals, which means they may be admitted to a different hospital than their original point of contact. For this study, we assume that there are no unreported cases. Asymptomatic cases fall within either stage $I$ or $H$. The model can be succinctly expressed using delay differential equations as follows:

$$
\begin{aligned}
\frac{dS}{dt} &= -S\frac{\beta_L\left(I+\sum_j\left(I_M^j+I_V^j\right)\right)}{N},\\
\frac{dE}{dt} &= S\frac{\beta_L\left(I+\sum_j\left(I_M^j+I_V^j\right)\right)}{N} - E\left(t-\tau_{E\to I}\right),\\
\frac{dI}{dt} &= E\left(t-\tau_{E\to I}\right) - I\left(t-\tau_{I\to H}\right),\\
\frac{dR}{dt} &= \sum_j Q^j(t-\tau_{Q\to R}).\\
\frac{dS_M^i}{dt} &= -S_M\left(\frac{\beta_{MM}I_M^i+\beta_{PM}(I_P^i+H^i)+\beta_{VM}I_v^i}{N^i}+\frac{\beta_L I}{N}\right),\\
&\text{for } i\in 1,2,\ldots,36,\\
\frac{dS_P^i}{dt} &= -S_P\frac{\beta_{MP}I_M^i+\beta_{PP}(I_P^i+H^i)+\beta_{VP}I_v^i}{N^i},\\
\frac{dS_V^i}{dt} &= -S_P\left(\frac{\beta_{MV}I_M^i+\beta_{PV}(I_P^i+H^i)+\beta_{VV}I_v^i}{N^i}+\frac{\beta_L I}{N}\right),\\
\frac{dE_M^i}{dt} &= S_M\left(\frac{\beta_{MM}I_M^i+\beta_{PM}(I_P^i+H^i)+\beta_{VM}I_v^i}{N^i}+\frac{\beta_L I}{N}\right) - E_M^i(t-\tau_{E\to I}),\\
\frac{dE_P^i}{dt} &= S_P\frac{\beta_{MP}I_M^i+\beta_{PP}(I_P^i+H^i)+\beta_{VP}I_v^i}{N^i} - E_P^i(t-\tau_{E\to I}),\\
\frac{dE_V^i}{dt} &= S_P\left(\frac{\beta_{MV}I_M^i+\beta_{PV}(I_P^i+H^i)+\beta_{VV}I_v^i}{N^i}+\frac{\beta_L I}{N}\right) - E_V^i(t-\tau_{E\to I}),\\
\frac{dI_M^i}{dt} &= E_M^i\left(t-\tau_{E\to I}\right) - I_M^i(t-\tau_{I\to Q}),\\
\frac{dI_P^i}{dt} &= E_P^i\left(t-\tau_{E\to I}\right) - I_P^i(t-\tau_{I\to Q}),\\
\frac{dI_V^i}{dt} &= E_M^i\left(t-\tau_{E\to I}\right) - I_V^i\left(t-\tau_{I\to Q}\right) - I_V^i(t-\tau_{I\to H}),\\
\frac{dI_M^i}{dt} &= E_M^i\left(t-\tau_{E\to I}\right) - I_M^i(t-\tau_{I\to Q}),\\
\frac{dI_P^i}{dt} &= E_P^i\left(t-\tau_{E\to I}\right) - I_P^i(t-\tau_{I\to Q}),\\
\frac{dI_V^i}{dt} &= E_M^i\left(t-\tau_{E\to I}\right) - I_V^i\left(t-\tau_{I\to H}\right),\\
\frac{dQ^i}{dt} &= I_M^i\left(t-\tau_{I\to Q}\right) + I_P^i\left(t-\tau_{I\to Q}\right) + I_V^i\left(t-\tau_{I\to Q}\right) - Q^i(t-\tau_{Q\to R}),\\
\frac{dH^i}{dt} &= \sum_j aI_V^j\left(t-\tau_{I\to H}\right) + aI\left(t-\tau_{I\to H}\right) - H^i\left(t-\tau_{H\to Q}\right).
\end{aligned}
$$

#### 2 Model simulation

This study focused on the Gangnam District in Seoul, Korea, known for its advanced medical infrastructure. The district comprises two tertiary referral hospitals (mean bed capacity: 1,533), two general hospitals (mean bed capacity: 225), and 32 smaller hospitals (mean bed capacity: 73) [2]. The population of the local community is 650,000 [3]. Notably, we excluded smaller clinics from our analysis. Our model simulation commenced with the introduction of a single primary case into the local community. To align with the Korean context in 2015, non-pharmaceutical interventions began six days after the admission of the index case, coinciding with outbreak recognition. These interventions restrict hospital visitors and reduce transmission rates within the hospital, resulting in a 41.02% reduction. Detailed methods for estimating transmission rates and this reduction are presented in the subsequent subsection. We employed the modified Gillespie algorithm for model simulation, running each setting 10,000 times [4].

### 3 Parameter estimation

Kim's study yielded valuable insights into individual hosts, considering their host type (medical staff, patients, visitors). The analysis encompassed anticipated exposure times, expected transmission times, isolation times, and the population size at Pyeongtaek St. Mary's Hospital PMH [5]. We denote the transmission rate as $\beta_{AB}$, where subscripts A and B correspond to infector and infectee types, respectively. Subscripts indicate host categories: medical staff, patients, and hospital visitors. Subsequently, we categorized individuals as either infected or uninfected. The group of infected individuals is denoted by $\Lambda_I$, while the uninfected group is represented as $\Lambda_S$. We introduced identifiers $D_i$ and $\tilde{D}_i$ to distinguish the host types within $\Lambda_S$ and $\Lambda_I$, respectively. Additionally, let $T$ denote the discrete time points during the outbreak. Given the count of infectors at a specific time $k$ denoted as $I_j(k)$, where subscript $j$ indicates the host type, we derive the likelihood of hosts in $\Lambda_S$ and $\Lambda_I$ as follows:

$$
\begin{aligned}
L_S &= \prod_{i\in\Lambda_S}\left\{\prod_{k\in T}\exp\left(\sum_{j\in\{M,P,V\}} -\beta_{jD_i}\frac{\mathbf{I}_j(k)}{N}\right)\right\},\\
L_I &= \prod_{i\in\Lambda_I}\left\{\prod_{k\in T}\left(1-\exp\left(\sum_{j\in\{M,P,V\}} -\beta_{j\tilde{D}_i}\frac{\mathbf{I}_j(k)}{N}\right)\right)\right\},\\
L(B) &= L_S \times L_I,\\
\text{where } B &= \{\beta_{MM},\beta_{MP},\beta_{MV},\beta_{PM},\beta_{PP},\beta_{PV},\beta_{VM},\beta_{VP},\beta_{VV}\}.
\end{aligned}
$$

Our objective is to determine the optimal value of $B$ that maximizes the likelihood function $L(B)$. Notably, we set transmission rates $\beta_{MM}$, $\beta_{MP}$, and $\beta_{MV}$ to zero based on epidemiological investigations, which revealed no contagious period involving medical staff at PMH. Additionally, we assumed symmetric transmission between patients and visitors, that is, $\beta_{PV} = \beta_{VP}$. To sample the parameters and estimate the distributions of transmission rates, we employed the Metropolis-Hastings algorithm [6]. Furthermore, we intentionally set the transmission rate in the local community ($\beta_L$) to 0.1, which is lower than the transmission rate within the hospital. This choice implies a basic reproduction number of approximately 0.5 in the local community. Notably, in Korea, only one infection case occurred outside the hospital among all reported cases [7].

The results of the transmission rate estimation are depicted in Fig 3. Panel A illustrates the transmission rates within the hospital, categorized by infector and infectee types. In Panel B, we present the reduction in transmission rates following outbreak recognition. Specifically, when the infector is a patient, the values are estimated as mean 0.16 (95% confidence interval (CI) [0.01, 0.47]), 1.40 (95% CI [0.84, 2.07]), and 0.49 (95% CI [0.26, 0.79]) when the infectee is medical staff, patient, and visitor, respectively. When the infector is a visitor, the values are estimated as mean 0.69 (95% CI [0.03, 1.88]), 0.59 (95% CI [0.02, 2.06]), and a0.15 (95% CI [0.00, 0.56]) when the infectee is medical staff, patient, and visitor, respectively. The reduction in the infection transmission rate after outbreak recognition is an average of 41.02% (95% CI [19.88, 72.28]). These estimated parameter values inform our simulation.

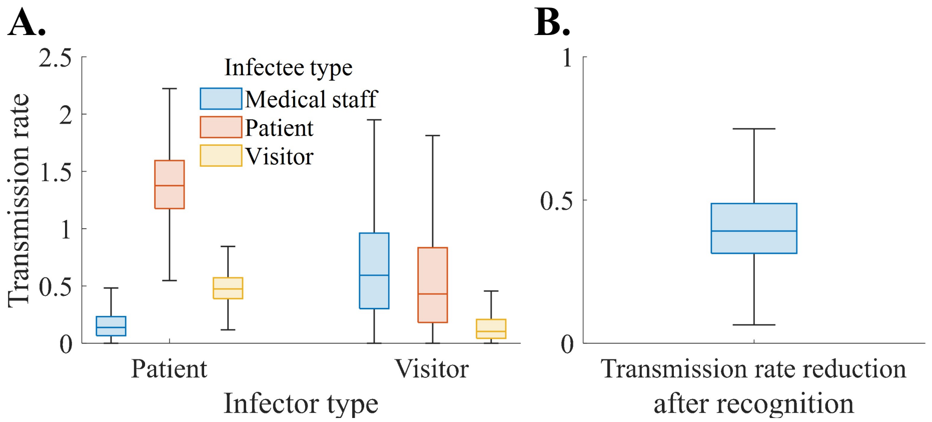


**Fig 3.** Estimated parameter distribution are depicted using box-and-whisker plots. Panel (A) illustrates the transmission rates within the hospital, while Panel(B) represents the reduction in transmission rates following outbreak recognition.

2. Nationwide status of hospitals and pharmacies [Internet], Health Insurance Review & Assessment Service. Available from: `https://opendata.hira.or.kr/op/opc/selectOpenData.do?sno=11925&publDataTpCd=&searchCnd=&searchWrd=%EC%A0%84%EA%B5%AD&pageIndex=1`

3. Population by administrative district (city, county, district) and gender [Internet]. Ministry of the Interior and Safety. Available from: `https://kosis.kr/statHtml/statHtml.do?orgId=101&tblId=DT_1B040A3`

4. Anderson DF. A modified next reaction method for simulating chemical systems with time dependent propensities and delays. The Journal of chemical physics. 2007;127(21).

5. Kim KM, Ki M, Cho SI, Sung M, Hong JK, Cheong HK, Kim JH, Lee SE, Lee C, Lee KJ, Park YS. Epidemiologic features of the first MERS outbreak in Korea: focus on Pyeongtaek St. Mary's Hospital. Epidemiology and health. 2015;37.

6. Chib S, Greenberg E. Understanding the metropolis-hastings algorithm. The american statistician. 1995;49(4):327-35.

7. Ki M. 2015 MERS outbreak in Korea: hospital-to-hospital transmission. Epidemiology and health. 2015;37.
